# Stable isotope labeling kinetics of neurofilament light *in vitro* and *in vivo*

**DOI:** 10.1101/2025.01.10.24319636

**Authors:** Claire A. Leckey, Tatiana A. Giovannucci, John B. Coulton, Yingxin He, Chihiro Sato, Nupur Ghoshal, Tharini Vignarajah, Zane Jaunmuktane, Nicolas R. Barthélemy, Henrik Zetterberg, Donald L. Elbert, Kevin Mills, Selina Wray, Randall J. Bateman, Ross W. Paterson

**Affiliations:** UCL Queen Square Institute of Neurology, London, United Kingdom; UCL Great Ormond Street Institute of Child Health, London, United Kingdom; Department of Neurology, Washington University School of Medicine, St. Louis, MO, USA; The Tracy Family SILQ Center, Washington University School of Medicine, St.Louis, MO, USA; Queen Square Brain Bank for Neurological Disorders, Department of Clinical and Movement Neurosciences, University College London; Clinical Neurochemistry Laboratory, Sahlgrenska University Hospital, Sweden, Mölndal, Sweden; Department of Psychiatry and Neurochemistry, Institute of Neuroscience and Physiology, the Sahlgrenska Academy at the University of Gothenburg, Mölndal, Sweden; UK Dementia Research Institute at UCL, London, UK; Hong Kong Center for Neurodegenerative Diseases, Clear Water Bay, Hong Kong, China; Wisconsin Alzheimer’s Disease Research Center, University of Wisconsin School of Medicine and Public Health, University of Wisconsin-Madison, Madison, WI, USA; Department of Neurology, University of Washington, Seattle, WA, USA; Dementia Research Centre, UCL Queen Square Institute of Neurology, London, United Kingdom; Darent Valley Hospital, Dartford, United Kingdom

**Author notes:** Corresponding author: Ross William Paterson, Address: 8-11 Queen Square London, WC1N 3AR. These authors contributed equally (Claire A. Leckey, Tatiana A. Giovannucci, Randall J. Bateman).

## Abstract

**Importance:** Neurofilament light (NfL) is elevated in CSF and blood across a range of traumatic, inflammatory and neurodegenerative diseases of the central nervous system, and has been increasingly included in clinical trials as an outcome measure of target engagement. Interpreting trajectories of NfL post-treatment has been challenging, prompting a greater need and focus on understanding its pathophysiology.

**Objective:** We measured NfL kinetics in the human central nervous system using stable isotope labeling kinetics (SILK).

**Design:** Observational study. Participants underwent SILK protocol. Infusion of 16 hours with 4mg/kg/h and follow-up lumbar punctures scheduled at 7, 14, 60 and 120 days post-labeling.

**Setting:** Referral center – specialist neurology clinic.

**Participants:** Participants with diagnosed primary tauopathies (n=10) were recruited to the Human CNS Tau Kinetics in Tauopathies (TANGLES) study. A control case was examined *post-mortem* to assess the technical background of the SILK method.

**Exposure:** Intravenous infusion of ^13^C_6_-leucine, with rates of label incorporation representative of fractional synthesis and fractional clearance rates *in vivo* and *in vitro*.

**Main outcome and Measure:** Level of incorporation of ^13^C_6_-leucine tracer into newly-translated NfL divided by the pool of NfL with previously incorporated ^12^C_6_-leucine, expressed as a percentage tracer-to-tracee (TTR) ratio.

**Results:** NfL is rapidly translated in human brain within hours but takes 53 – 162 days to appear in cerebrospinal fluid (CSF). Labeled NfL remains detectable in *post-mortem* brain tissue 1.5 years post-labeling, indicating an extremely slow turnover in the human CNS. Together, these data suggest the greatest contribution of CSF NfL in neurodegeneration is from slow release of a large pool of previously translated NfL. However, release of newly translated NfL makes a significant contribution.

**Conclusion and relevance:** Rapid increases in CSF NfL seen within weeks of disease processes or interventions are likely to reflect release of pre-existing NfL from damaged neurons, but later increases in NfL (>3 months) may also reflect new NfL translation and release. Clinical trials using NfL as an outcome measure to track neurodegeneration would particularly benefit from substantially longer follow-up periods due to the slow turnover of the protein in the central nervous system.

## Introduction

Neurofilament light chain (NfL) has emerged as a promising biomarker of neurodegeneration in blood and cerebrospinal fluid (CSF)^1^. Elevated NfL has been observed across a wide range of neurological conditions^2,3^. Remarkably, NfL levels dynamically reflect acute neural damage, rapidly increasing within 0 – 48 hours following traumatic brain injury (TBI) and hypoxic brain injury^4,5^, and acutely following clinical relapse in multiple sclerosis (MS)^6^. It can also fall in response to clinically successful disease-modification^7,8^. This dynamic behavior makes NfL an attractive biomarker for disease monitoring and prognostication.

Yet, precise biological implications of elevated NfL levels remain unclear. NfL is a neuroaxonal protein that assembles into heteropolymers in the central nervous system (CNS) to form the filament network that sustains axonal architecture^9^, the distribution of organelles across axons^10^ and the stability of synaptic receptors^11–13^. Possible explanations for the appearance of NfL in CSF include unregulated passive release from disrupted axons, active release through increased processing and secretion of NfL into the three major proteoforms^14–16^, upregulated NfL synthesis, or a combination of these mechanisms.

Determining the timing and magnitude of NfL responses is important for understanding when target engagement occurs and how to interpret changes in NfL levels during clinical trials. Furthermore, the disappearance of NfL from CSF is also likely to be under the influence of several possible clearance mechanisms and routes, which may be disrupted in neurodegeneration or by the intrathecal delivery of therapies.

To address these open questions, we developed a Stable Isotope Labeling Kinetics (SILK) assay for NfL, a technique previously used to determine the kinetics of proteins related to neurodegeneration^17–19^. NfL-SILK can be used to measure NfL kinetics in CSF and estimate the timing of NfL synthesis and release in humans.

## Methods

### Human CNS Tau Kinetics in Tauopathies (TANGLES) study design

The TANGLES study is a project between Washington University in St Louis and University College London (UCL) aiming to quantitate tau kinetics in patients with primary tauopathies. For this study, CSF samples from study participants recruited and labeled at UCL were used to determine NfL turnover *in vivo*.

The UCL TANGLES study was approved by the London-Bloomsbury ethic committee (reference: 18/LO/8601). Criteria for recruitment is included in the Supplementary Methods.

### Differentiation of iPSC into cortical neurons

Ctrl1 and Ctrl2 refer to the well characterized SIGi1001-a-1 and RBi001-a, respectively, both available via Sigma Aldrich. Ctrl3 refers to a patient-derived cell line kindly shared by Dr Tilo Kunath^20^.

iPSCs were differentiated to cortical neurons using established protocols^21,22^. We characterized all cell lines for neuronal developmental markers by quantitative PCR and immunocytochemistry (see Supplementary Methods and Supplementary Tables 1-3).

### NfL-SILK in iPSC-derived neurons

iPSC-derived neurons at 70 days-*in-vitro* (DIV) were labeled with N2B27 media containing equal amounts of ^13^C_6_-leucine (50 mol, 100% tracer-to-tracee ratio [TTR] media) for 24 days (full media change every three days), followed by culture in label-free N2B27 for 15 – 21 days. NfL-SILK sample processing details are reported in Supplementary Methods.

### NfL-SILK in human subjects

#### UCL TANGLES SILK study

Participants were labeled as previously described^18^, but with some site-specific adaptations (see Supplementary Methods).

#### UCL Normal Pressure Hydrocephalus SILK study

The UCL NPH SILK study was approved by the Bloomsbury ethics committee and all individuals provided informed written consent. Recruitment criteria and labeling protocol are described in the Supplementary Methods.

#### IP-MS/MS of NfL in CSF

To measure ^13^C_6_-leucine label incorporation into NfL *in vivo*, CSF was prepared and analyzed by peptide IP-MS/MS as previously described^15^, but with the following adaptations: CSF sample volume increased to 1000 µL prior to spiking with 1 ng of a heavy labeled [^13^C, ^15^N -Arg/Lys] recombinant NfL standard (Promise Proteomics, France).

#### IP-MS/MS of NfL from soluble and insoluble fractions of post-mortem brain tissue

Brain samples from frontal cortex were processed following protocols from Mukherjee *et al.*^23^, briefly described in the Supplementary Methods.

For immunoprecipitation of NfL from the insoluble fraction, lyophilized proteins were resuspended in a small amount (ca 20 µl) of 70% FA and neutralized with Tris-HCl pH 11. Sarkosyl soluble and insoluble fractions at 1 mg/ml in 500 µl were used for IP, adding Triton-X100 to a final concentration of 0.1%. Immunoprecipitation is further described in “Extracellular NfL-SILK” (see Supplementary Methods).

### NfL-SILK Quantitation

Ultra performance liquid chromatography-tandem mass spectrometry (UPLC-MS/MS) analysis was performed as previously described^19^, but with minor adaptions to monitor SILK labeled peptides (see Supplementary Table 4). Acquired data was imported into Skyline software (MacCoss Lab, University of Washington). Extracted peak areas were used to calculate the tracer-to-tracee ratio (TTR), fractional synthesis rate (FSR), fractional clearance rate (FCR) and half-life of NfL following methods described in the Supplementary Methods section.

### Statistics

Statistical analyses were performed using GraphPad Prism v10.1.2. To test for Gaussian distribution, the Shapiro-Wilk normality test was used. If the normality test was passed, data were analyzed by Student’s unpaired t-test (two groups) or by ANOVA, otherwise. If the data were not normally distributed, statistical analysis was performed using the nonparametric Mann–Whitney test (two groups) or Kruskal–Wallis test for multiple comparisons, with Dunnett’s or Tukey’s test to adjust for multiple comparisons.

## Results

### NfL profiling in brain, CSF and neurons

We first employed a mass spectrometry assay together with immunoprecipitation (IP) to characterize NfL, including potential truncated species, in brain and CSF from a cohort of individuals with primary tauopathies. NfL was immunopurified from lumbar CSF (n=10), as well as from three brain samples from the same cohort and a non-degenerative case donated *post-mortem*. Brain samples were biochemically fractionated into sarkosyl soluble and insoluble fractions to determine the solubility profile of NfL^24^. Details of the cohort can be found in **Table 1**.

**Table 1.**
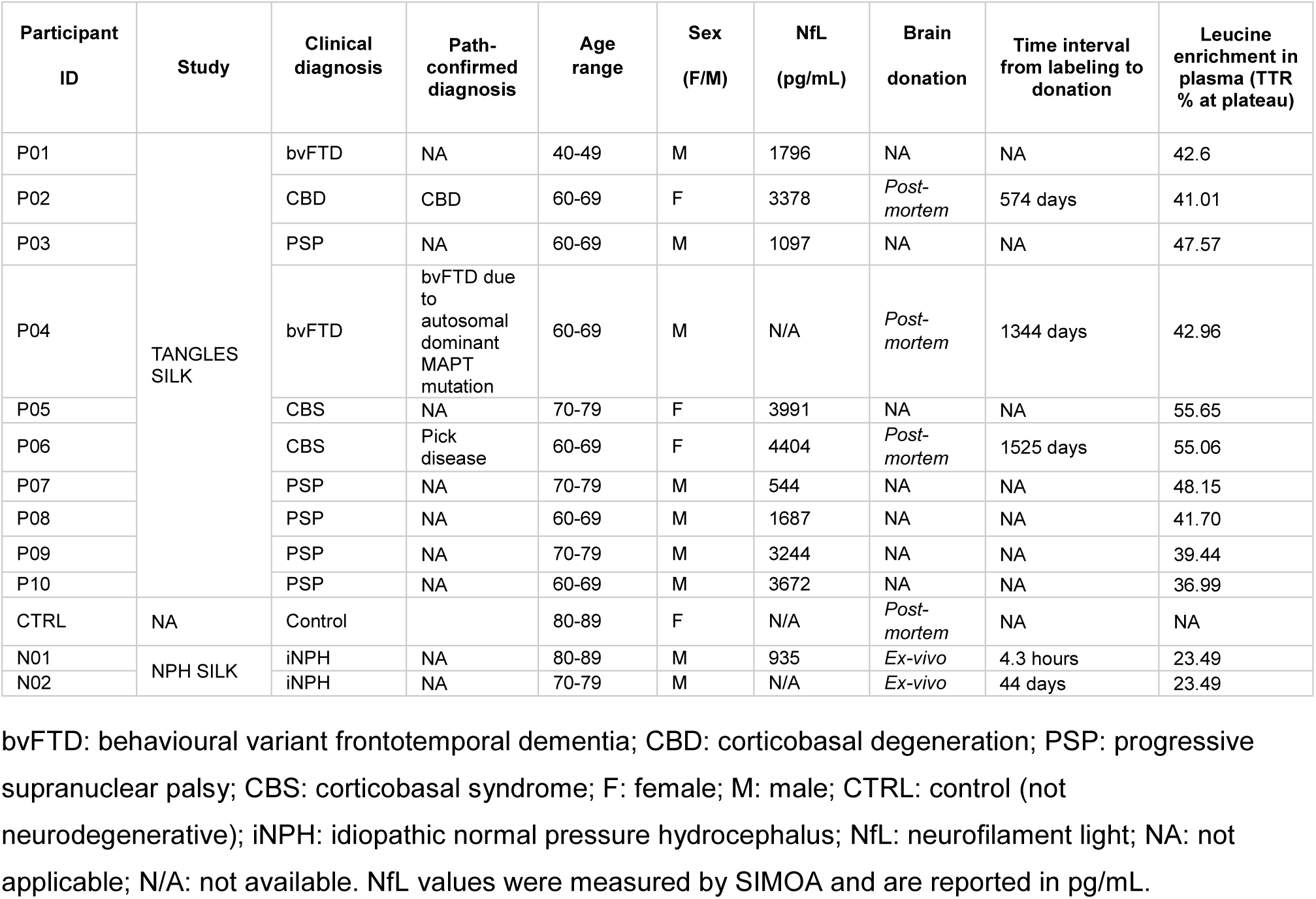
Cohort characteristics.

We used a combination of three custom monoclonal antibodies targeting discrete regions across the NfL sequence (**Figure 1A**). We normalized peptide abundances to the Coil 2b peptide (residues [324-331]) to aid comparisons between compartments. The predominant form of NfL in CSF consisted of proteoforms containing mostly the Coil 2b domain, whilst brain samples contained higher relative abundances of peptides in the head, Coil 1a and Coil 1b domains (**Figure 1B,C**; **Suppl. Figure 1**). These results are consistent with previous studies describing full length NfL recovery in brain but truncated NfL forms in CSF using the same methods^14,15^. We found that NfL was most abundant in the soluble brain fraction but was also recovered in smaller amounts in the insoluble fractions, largely maintaining the same peptide profile (**Figure 1B**; **Suppl. Figure 1**). Comparison to a cognitively unimpaired brain showed that the proportion of NfL in the insoluble fraction was 4.2-fold higher in P02 and 2.5-2.2-fold higher in P04 and P06 (**Suppl. Figure 1**). This is in line with previous mass spectrometry-based reports identifying NfL peptides in insoluble protein inclusions and neurofibrillary tangles by mass spectrometry^25,26^. The N-terminal domain of NfL is known to be a region of low complexity and a target of post-translational modifications including phosphorylation and glycosylation^27^. Our assay included a peptide in the N-terminal domain of NfL (residues [31-37]), which was recovered from brain but largely reduced in CSF (**Figure 1B,C**), and was more abundant in the control than in the tauopathy brains (**Suppl. Figure 1**).

**Figure 1.**
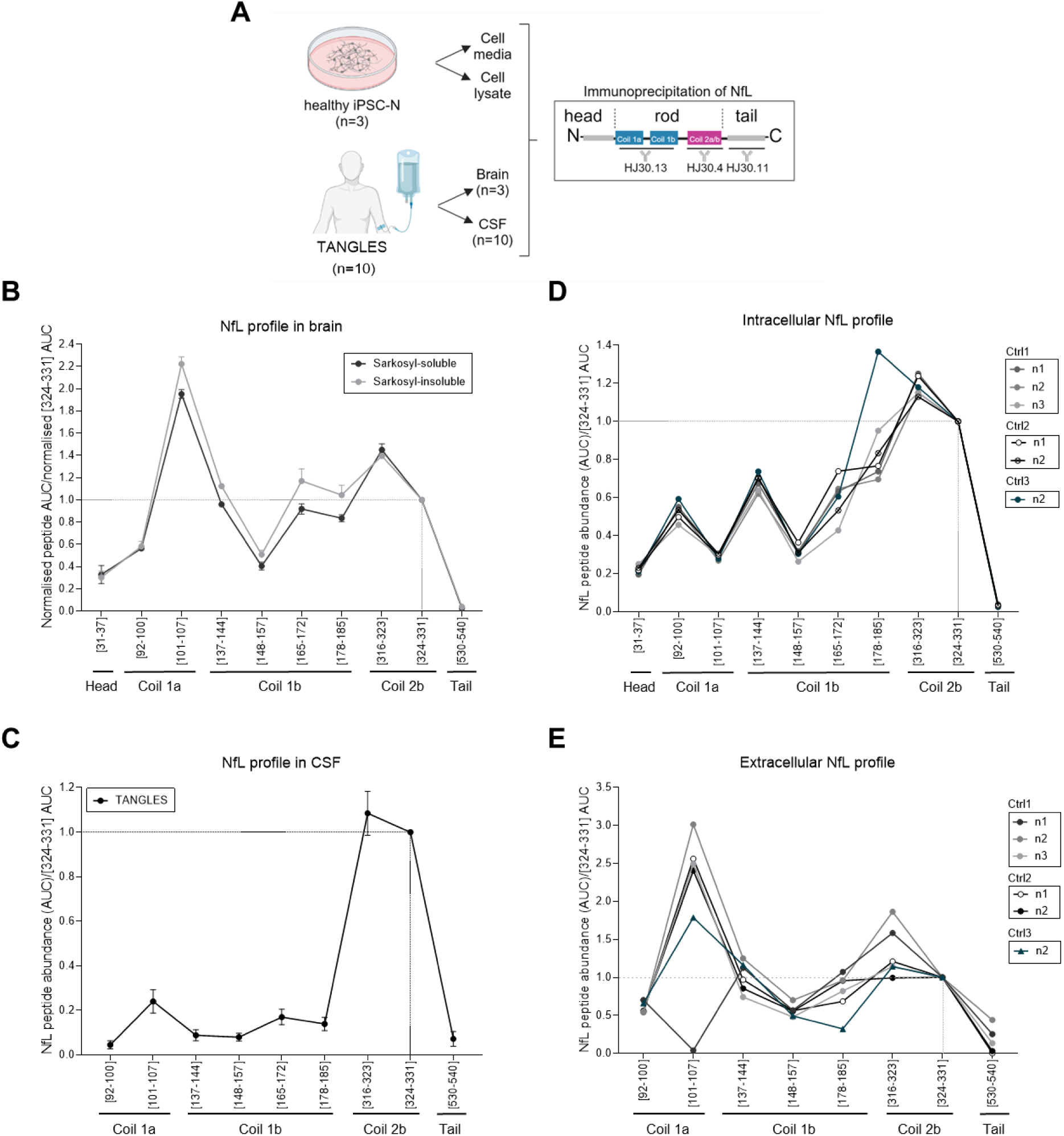
NfL profiling *in vivo* and *in vitro*. **A)** Schematic of samples and processing. **B)** Brain recovery of NfL proteotypic peptides. Each peptide is indicated by its first-and last position in the canonical NfL amino acid sequence between brackets. *Post-mortem* cortical samples were processed into sarkosyl-soluble and insoluble fractions. Data shown as mean ± SD (n=3). **C)** CSF recovery of NfL proteotypic peptides. Data shown as mean ± SD (n=10). **D)** Intracellular recovery of NfL in iPSC-derived neurons from three control lines. **E)** Extracellular recovery of NfL in conditioned media from iPSC-derived neurons from three control lines. Independent inductions shown (n=3 Ctrl1; n=2 Ctrl2; n=1 Ctrl3). All peptide profiles are shown as abundance (area under the curve [AUC]) relative to peptide [324-331], indicated by dashed lines.

Neurons from human induced-pluripotent stem cells (iPSC) are a translational model uniquely placed to examine neural function and model disease processes *in vitro*. We first tested how the NfL profile recovered from cell extracts and conditioned media compared to intra-and extracellular compartments in humans (cell lysate to brain, and conditioned media to CSF, respectively). Three iPSC lines derived from cognitively normal controls at the time of biopsy were differentiated into cortical neurons^22^ in several experimentally independent batches. We found the N-terminal peptide (residues [31-37]) in cell lysates and brain, which was largely reduced in cell media and CSF. Contrary to CSF but similar to brain, cell-conditioned media was rich in Coil 1a peptides (residues [101-107]). The recovery of peptides from the C-terminal tail domain was low in all compartments. Coil 2b peptides, the most abundant in CSF across neurodegenerative conditions^15^, and the most widely measured part of the protein as a clinical and research biomarker, were recovered from all compartments *in vivo* and *in vitro*.

### NfL kinetics in induced pluripotent stem cell (iPSC)-derived cortical neurons

To study NfL kinetics *in vitro*, we next adapted the mass spectrometry method for the detection of isotopically-labeled leucine incorporation into six proteotypic peptides (namely, unique to NfL) following a SILK paradigm (**Suppl. Table 4**) using three iPSC lines derived from non-degenerative controls at the time of biopsy (**Suppl. Table 1**). Expression of pluripotency markers in iPSCs, as well as expression of neuronal markers by immunostaining and quantitative PCR demonstrated equivalent differentiation into cortical neurons across lines and inductions (**Suppl. Figure 2**). At 100 DIV, all lines expressed equivalent levels of the neuronal-specific tubulin *TUBB3*, whilst *NEFL* expression differed between lines, with Ctrl1 showing the lowest expression levels (**Suppl. Figure 2**).

NfL intracellular turnover was relatively slow in all three lines (**Figure 2B,C,D** and **Suppl. Figure 3**), with a similar half-life between control lines (5.06 ± 0.96 days in Ctrl1 cells; 6.95 ± 2.79 days in Ctrl2; see **Table 2**). NfL had similar kinetics to microtubule-associated protein tau and comparatively much slower than the fast amyloid precursor protein (**Suppl. Figure 3**). Cell membrane integrity remained stable during the experiment (**Suppl. Figure 3**). There were no differences between the individual peptide’s half-lives (one-way ANOVA, F (5,13) =1.664, p=0.2124 with Tukey’s multiple comparisons test, ns) (**Suppl. Figure 4**). For all the peptides monitored, the resulting extracellular turnover curve was shifted, resulting in a delay in appearance of labeled NfL reflected in a delay of 3 to 6 days to achieve maximum labeling (“time-to-peak", a delay of 3 days in Ctrl1 cells, and of 6 days in Ctrl2 and Ctrl3). This is consistent with previous findings in tau SILK^18^ and might be the result of the transport from intra-to extracellular and suggests it would not only originate from dying neurons at any given timepoint (where no delay would be expected). This is also supported by the differences in relative abundance of Coil 1a and Coil 2b peptides between lysates and media (**Figure 1D,E**). Clearance rates were much higher in cell lysates (mean Fractional Clearance Rate [FCR] 3.67 ± 0.51) than media (mean FCR 1.92 ± 0.48), consistent with a lack of clearance mechanisms from the media in this 2D-*in vitro* system and suggesting that there is limited degradation/proteolysis of NfL in neuronal conditioned media (**Suppl. Table 2**).

**Figure 2.**
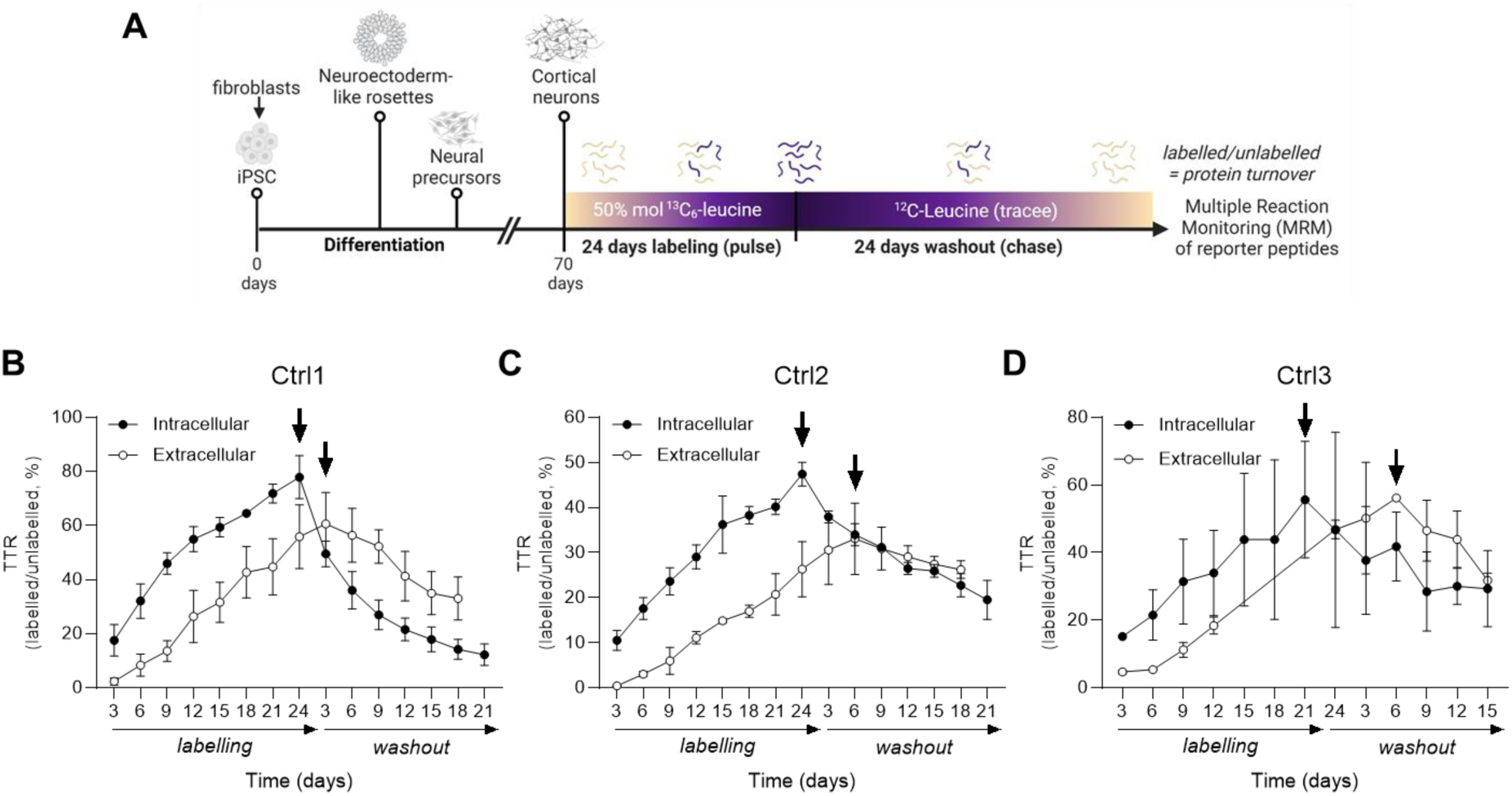
NfL kinetics in iPSC-derived neurons from three non-degenerative donors. **A)** Schematic. **B)** Ctrl1 results (n=3-4 independent inductions). **C)** Ctrl2 results (n=2 independent inductions). **D)** Ctrl3 intracellular results (n= 2 independent inductions). Datapoints represent the mean tracer-to-tracee ratio (TTR) of all peptides at any given timepoint from all inductions ± SD. Indicated with arrows is the time-to-peak of each intra-and extracellular kinetic curves.

**Table 2.** Kinetic measurements in iPSC-derived neuron cell lysates.

| Half-life |  |  |  |  |  |  |  |  |  |  |  |  |
| --- | --- | --- | --- | --- | --- | --- | --- | --- | --- | --- | --- | --- |
|  |  | Ctrl1 |  |  |  |  | Ctrl2 |  |  | Ctrl3 |  |  |
| Peptide | Amino acids | n1 | n2 | n3 | n4 | Mean half-life (d) ± SD | n1 | n2 | Mean half-life (d) | n1 | n2 | Mean half-life (d) |
| AQLQDLNDR | 92-100 | 4.48 | 6.51 | 5.07 | 7.10 | 5.79 ± 1.22 | 9.11* | 4.44 | 6.78 | NC | NC | NC |
| ALYEQEIR | 137-144 | 2.75 | 4.99 | 3.42 | 4.87 | 4.01 ± 1.10 | 70.45* | 6.12 | 38.29 | 15.71* | 2.33* | NC |
| LAAEDATNEK | 148-157 | 2.34 | 4.91 | 3.17 | 5.20 | 3.91 ± 1.37 | 26.13* | 10.56 | 18.35 | 7.67 | 3.90* | 7.76 |
| FTVLTESAAK | 284-293 | NC | 5.42 | NC | 5.54 | 5.48 ± 0.08 | 10.35 | 7.73* | 9.04 | NC | NC | NC |
| TLEIEACR | 316-323 | 3.73 | 8.48 | NC | 6.54 | 6.25 ± 2.39 | 25.37* | 5.68 | 15.53 | NC | NC | NC |
| GMNEALEK | 324-331 | NC | 4.50 | NC | 5.37 | 4.93 ± 0.62 | 5.29 | 3.78 | 4.53 | NC | NC | NC |
| | Mean all | | | | | 5.06 ± 0.96 | | | 6.95 ± 2.79 | | | 7.76 |
| Fractional Synthesis Rate (Intracellular) |  |  |  |  |  |  |  |  |  |  |  |  |
| Peptide | Amino acids | n1 | n2 | n3 | n4 | Mean FSR (%/d) ± SD | n1 | n2 | Mean FSR (%/d) ± SD | n1 | n2 | Mean FSR (%/d) ± SD |
| AQLQDLNDR | 92-100 | 3.02 | 1.78 | 2.01 | 1.83 | 2.16 ± 0.58 | 1.38 | 1.37 | 1.37 ± 1.48 | NC | NC | NC |
| ALYEQEIR | 137-144 | 4.55 | 3.00 | 3.30 | 2.82 | 3.42 ± 0.78 | 1.84 | 1.71 | 1.77 ± 1.65 | 2.52 | 2.23 | 2.38 ± 0.21 |
| LAAEDATNEK | 148-157 | 5.25 | 3.26 | 3.27 | 2.76 | 3.63 ± 1.10 | 1.86 | 1.62 | 1.74 ± 1.22 | 2.58 | 2.17 | 2.37 ± 0.00 |
| FTVLTESAAK | 284-293 | NC | 2.93 | NC | 2.79 | 2.86 ± 0.10 | 2.20 | 1.87 | 2.03 ± 2.06 | NC | NC | NC |
| TLEIEACR | 316-323 | 4.79 | 2.09 | NC | 2.36 | 3.08 ± 1.49 | 1.80 | 1.60 | 1.70 ± 1.30 | NC | NC | NC |
| GMNEALEK | 324-331 | NC | 3.13 | NC | 1.89 | 2.51 ± 0.88 | 1.92 | 1.89 | 1.91 ± 1.41 | NC | NC | NC |
| | Mean all | | | | | 2.94 ± 0.55 | | | 1.75 ± 0.20 | | | 2.37 ± 0.1 |
| Fractional Clearance Rate (Intracellular) |  |  |  |  |  |  |  |  |  |  |  |  |
| Peptide | Amino acids | n1 | n2 | n3 | n4 | Mean FCR (%/d) ± SD | n1 | n2 | Mean FCR (%/d) ± SD | n1 | n2 | Mean FCR (%/d) ± SD |
| AQLQDLNDR | 92-100 | 12.12 | 7.28 | 8.39 | 8.10 | 8.97 ± 2.15 | 4.56 | 2.46 | 3.51 ± 1.48 | NC | NC | NC |
| ALYEQEIR | 137-144 | 10.60 | 7.04 | 7.96 | 7.17 | 8.19 ± 1.66 | 5.10 | 2.76 | 3.93 ± 1.65 | 8.24 | 10.37 | 9.31 ± 1.51 |
| LAAEDATNEK | 148-157 | 7.74 | 7.14 | 8.18 | 7.69 | 7.69 ± 0.43 | 4.74 | 3.02 | 3.88 ± 1.22 | 7.96 | 8.34 | 8.15 ± 0.27 |
| **Fractional Clearance Rate (Intracellular) (continued).** |  |  |  |  |  |  |  |  |  |  |  |  |
|  |  | **Ctrl1** |  |  |  |  | **Ctrl2** |  |  | **Ctrl3** |  |  |
| Peptide | Amino acids | n1 | n2 | n3 | n4 | Mean FCR (%/d) ± SD | n1 | n2 | Mean FCR (%/d) ± SD | n2 | n3 | Mean FCR (%/d) ± SD |
| FTVLTESAAK | 284-293 | NC | 7.37 | NC | 7.87 | 7.62 ± 0.35 | 5.85 | 2.93 | 4.39 ± 2.06 | NC | NC | NC |
| TLEIEACR | 316-323 | 9.35 | 7.06 | NC | 7.27 | 7.89 ± 1.27 | 4.35 | 2.51 | 3.43 ± 1.30 | NC | NC | NC |
| GMNEALEK | 324-331 | NC | 5.95 | NC | 5.51 | 5.73 ± 0.31 | 3.89 | 1.89 | 2.89 ± 1.41 | NC | NC | NC |
|  | Mean all |  |  |  |  | 7.68 ± 1.08 |  |  | 3.67 ± 0.51 |  |  | 8.73 ± 0.82 |

SD: standard deviation; FSR: fractional synthesis rate (represented as percentage per day [d] ± SD); FCR: fractional clearance rate (represented as percentage per day [d] ± SD); n: induction number; amino acids in the consensus sequence of NfL as per Uniprot entry P07196; NC: not captured (peaks did not pass QC). The half-life estimates were calculated fitting the data into a one-phase decay model. Half-life measurements marked with an asterisk (\*) were deemed unreliable (the model could not report a double-sided confidence interval) and were not included in the calculation of the average.

### NfL kinetics in humans

To capture NfL dynamics *in vivo* we analyzed CSF and donated brain tissue from study participants from two SILK cohorts (**Table 2**); TANGLES (CSF and *post-mortem* brain in primary tauopathies) and Normal Pressure Hydrocephalus (NPH) SILK (*ex vivo* brain tissue from NPH patients). Having established a greater enrichment of Coil 2b peptides in TANGLES CSF during profiling (**Figure 1C**), label incorporation into NfL was measured using a peptide IP-MS/MS approach for Coil 2b peptide TLEIEACR (**Figure 3A**), while a full protein IP-MS/MS approach was used for brain tissue (**Figure 3A**) due to its more uniform NfL profile distribution across the protein’s structural domains (**Figure 1B**).

**Figure 3.**
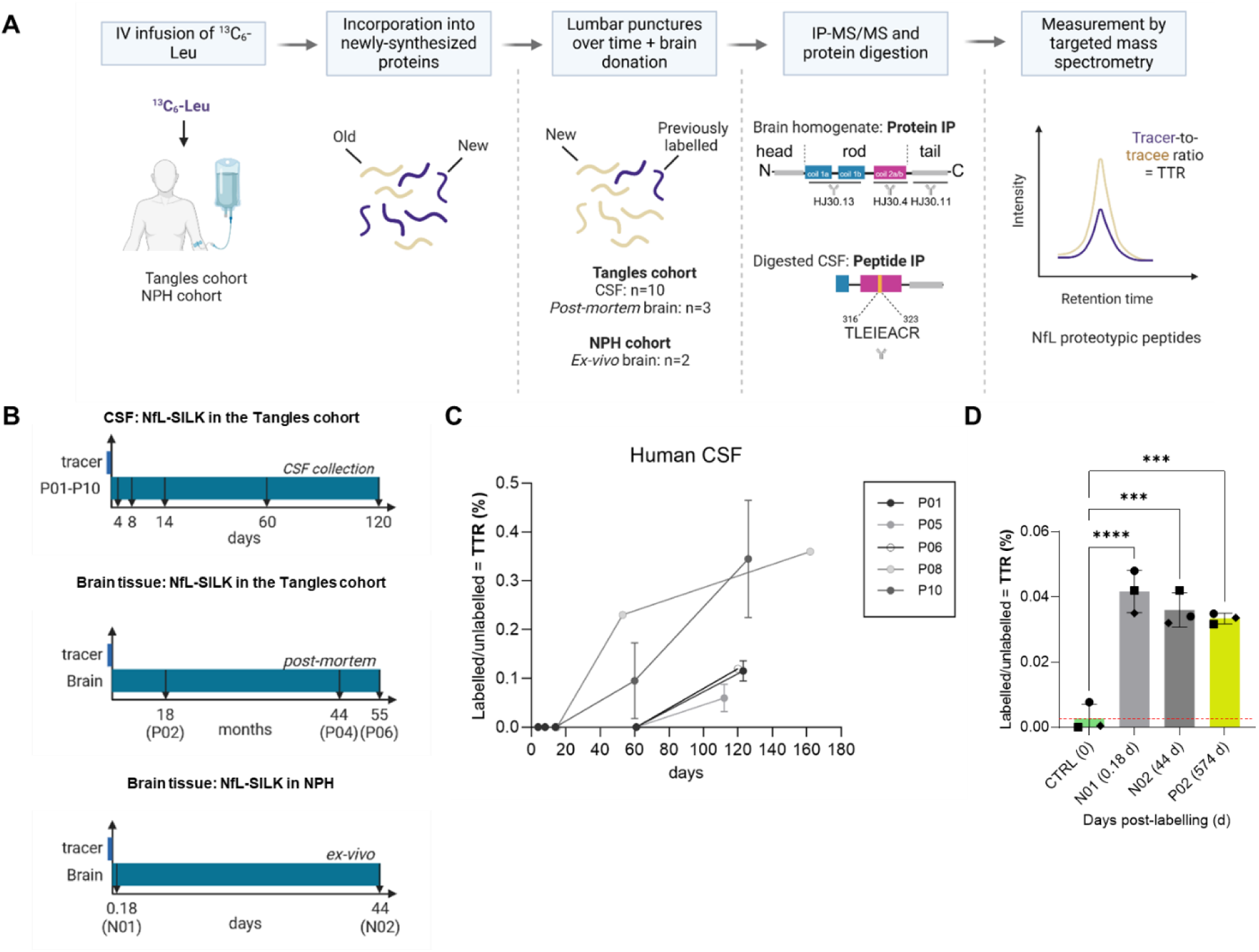
NfL kinetics in the human CNS. **A)** Overview of the NfL SILK analysis pipeline *in vivo*. **B)** NfL SILK labeling and collection protocol relevant to this study for CSF and brain in the TANGLES and NPH SILK studies. **C)** NfL kinetic curves in CSF from TANGLES participants. **D)** ^13^C6-leucine incorporation into NfL in brain tissue. Labeled NfL in brain tissue donated at 4 hours (N01), 44 days (N02) and 18 months (P02) compared to baseline abundance of ^13^C6-labeled NfL in a non-labeled control brain. Data are depicted as mean ± SD of three NfL proteotypic peptides: [148-157], depicted with a squared symbol; [178-185], depicted with a diamond-shaped symbol; and [324-331], depicted with a round symbol. One-way ANOVA with Tukey’s multiple comparison’s test. ***P ≤ 0.001; ****P ≤ 0.0001. Not shown = not significant. IV = intravenous.

The TANGLES cohort was selected for CSF SILK analysis due to the study’s longer pulse (16 hours 4mg/kg/hr ^13^C_6_-leucine infusion) and chase (up to 162 days) periods, making it well-suited for monitoring proteins with potentially slow turnover (**Figure 3B**). Labeled NfL was detected in 5/9 (56%) TANGLES study participants, with plotted kinetic curves suggesting NfL turnover to be very slow, with low label incorporation (0.04 – 0.36%) observed by the end of the chase (**Figure 3C**).

For determining NfL turnover in brain, *ex vivo* tissue was collected during insertion of ventriculoperitoneal shunting in two individuals with suspected NPH at 4.3 hours and 44 days post-labeling, while donated *post-mortem* tissue was analyzed at substantially longer timepoints post-labeling (18 – 55 months) from the TANGLES cohort (**Figure 3D**). Brain tissue was homogenized and fractionated, before NfL was enriched by protein IP-MS/MS to quantitate labeled NfL ratios in sarkosyl-soluble and sarkosyl-insoluble fractions. Due to the ethical unfeasibility of obtaining brain tissue from a single individual at multiple timepoints for a kinetic curve, NfL SILK data in brain tissue from three individuals were plotted as NfL TTR (in the detergent-soluble fraction) *vs* time of collection post-labeling. Combined, the data shows label incorporation into NfL at 0.03 – 0.05%, with a small, non-significant reduction in NfL TTR captured by 574 days post-label from both cohorts, which was the longest timepoint that could be reliably analyzed (**Figure 3D**). All labeled samples showed significantly higher incorporation of ^13^C_6_-leucine compared to a non-labeled brain, representative of the isotopic natural abundance level. Overall, the data indicate that NfL translation in the brain is detectable within hours of tracer infusion, and that newly synthesized NfL remains metabolically stable over months in the population studied.

## Discussion

We provide the first quantitation of NfL kinetics in the CNS and human neurons using SILK. We show that intracellular NfL translation occurs within 4 hours in *ex vivo* human brain, but detection of labeled (new) NfL in CSF is first observed around 53 days post-labeling. Together, this data suggests that NfL is translated rapidly in brain, while its release into CSF is very slow, with no peak of label incorporation or clearance captured during the study’s 5.4-month chase period. Labeled (new) NfL in brain can be detected at stable levels in cortical brain tissue as long as 1.5 years after SILK labeling. This is consistent with previous data in animal models suggesting there is a relatively small finite source of stable NfL in the human CNS and turnover of NfL is extremely slow^28,35,36^.

To study brain-CSF NfL dynamics, we analyzed CSF samples from the TANGLES cohort, and donated *post-mortem* brain tissue from three TANGLES participants, together with *ex vivo* tissue from two participants of the NPH SILK cohort.

In brain, fractionation and subsequent profiling determined NfL to be most abundant in the soluble fraction, and NfL present in the insoluble fractions of the brain to be higher in brains donated earlier than others, suggesting individuals with greater disease progression/severity may have more NfL within insoluble protein inclusions. This is supported by previous research of aggregated neurofilaments in neurodegenerative pathologies including AD, Parkinson disease, frontotemporal dementia and ALS^29–32^. We cannot exclude different NfL solubility profiles and/or regional differences between the different primary tauopathies analyzed, which future studies should address.

Analysis of labeled NfL in frontal cortex samples showed that NfL is rapidly translated, within hours of labeling, but remains in the stable cytoskeletal lattice from hours to months. Labeled NfL levels in sarkosyl-soluble fractions from *ex vivo* tissue samples taken at 4.3 hours and 44 days post-labeling (0.042 – 0.033% TTR) and at 574 days post-labeling in *post-mortem* brain tissue (0.033%) remained stable. Without access to samples between 44 – 574 days, we cannot discard dynamic changes during that period – however, the stable retention of newly-synthesized NfL within neurons is supported by the delayed detection of labeled NfL in CSF; first detected between 53 – 162 days post-labeling, and with only the start of the kinetic curve captured during the 5.4-month study period.

In humans, dynamic responses of NfL have been studied longitudinally by measuring changes in static CSF, plasma and/or serum NfL concentrations. This has been particularly instructive in scenarios where the steady state of NfL is challenged, e.g. acute brain injury (TBI, neuroinflammation or stroke). In TBI, rises are seen in CSF and plasma within 7 – 10 days, and fall to normal background levels within 120 – 180 days^4,33^. This has been interpreted as reflecting passive release of established reservoirs of axonal NfL rather than reflecting new NfL synthesis. We do not have access to SILK labeled individuals undergoing acute brain injury, but the timing of NfL appearance in our cohort supports this conclusion.

Clinical trials of disease-modifying therapies have brought particular sharp focus on the interpretation of NfL, particularly when used as a biomarker of therapeutic effect. Overall, NfL response across AD, HD and ALS trials has been mixed^8,34–39^, but it is likely to be relevant that the most successful clinical outcomes, e.g. SOD1 ALS trial, are associated with early significant reductions in NfL. Successfully interrupting neurodegeneration reduces the CSF and plasma pool of NfL which, given the time taken to translate and release NfL into the extracellular space we observed *in vivo*, is more likely to be explained by a reduction in NfL passive release rather than a downregulation of NfL synthesis.

Conversely, our data shows that newly translated NfL takes at least 53 – 112 days to appear in CSF, therefore NfL values measured after ∼4 months following a therapeutic intervention could reflect a contribution from NfL passive release (neurodegeneration or physiological axonal remodeling) *and/or* contributions from newly translated NfL. The biological relevance of the appearance of newly labeled NfL in CSF is uncertain, but could represent biological recovery or neuroregeneration, and highlights that the pool of NfL in CSF is more dynamic than previously appreciated. Further dynamic labeling studies are going to be critical to understand the relative contribution of passively released versus newly generated NfL across disease states, particularly when the steady state is disrupted through therapeutic intervention. Clinical labeling protocols reflecting the very long turnover of NfL, with long follow-up periods of a year or more and higher label quantity, will be required to fully capture NfL kinetics.

Studying NfL kinetics in human neurons *in vitro* provided evidence of rapid NfL translation and its delayed release to the extracellular space, which mirrored the *in vivo* findings but at an accelerated rate. While *in vivo* kinetics of NfL were found to be slower compared to tau in the human CNS (half-life of 23 ± 6.4 days)^18^, *in vitro* kinetics of intracellular NfL, with half-lives of 5.06 ± 0.96 days (Ctrl1) and 6.95 ± 2.79 days (Ctrl2), were similar to tau (6.74 ± 0.45 days)^18^. The faster turnover rate of NfL and tau observed *in vitro* compared to in *vivo* might be due to faster axonal remodeling, a faster metabolic rate *in vitro* and/or due to more resilient proteostasis mechanisms in the fetal-like phenotype of the iPSC-derived neuronal models used in both studies. Ultimately, NfL-SILK *in vitro* supported that intracellular protein kinetic events in the brain can be inferred from extracellular compartments.

This study has limitations. The reason for the differences in NfL peptide profiles between *in vitro* and *in vivo*, which could be developmental and change in more mature neurons, were not addressed. In the Ctrl1 line, the low *NEFL* expression levels coupled with the technical limitations of the NfL SILK assay resulted in a limited data set for this line. The clinical research labeling SILK protocol was not long enough to capture the maximum TTR of NfL, so further studies will be required to ascertain the full kinetic curve of NfL. Secondly, not all participants had evidence of NfL labeling during the 120 day follow up period (3/8). Since the limit of detection of labeled NfL was close to our measured values (**Suppl. Figure 5**), we cannot ascertain whether this is a technical limitation, or if it reflects an inability of neurons to generate new NfL due to more advanced neurodegenerative disease and neuronal loss. Notably, 3/5 of the individuals were labeling could be measured, died of their neurodegenerative disease within 24-months of participating in the study, reflecting a more advanced disease stage.

In summary, we describe a novel method for quantitating the kinetics of NfL *in vitro* and *in vivo*. We show that NfL is rapidly translated in human brain but takes 2 – 3 months before appearing in human CSF. NfL is likely to have a very long half-life in the human CNS.

## Data Availability

All data produced in the present study are available upon reasonable request to the authors.

## Acknowledgements

We are grateful to the research participants who participated in this study, to the Leonard Wolfson Experimental Neurology Centre at UCL for supporting sample collection, to the UCL Queen Square Brain Bank, to David M. Holtzman for kindly sharing the custom-made anti-NfL antibodies for protein IP used in this study and to all the members of The Neurofilament Light Consortium, specially Holly Soares, Omar Mabrouk, Antony Bannon and Ramakrishna Boyanapalli, for constructive feedback.

## Data availability

Data are available from the corresponding author on reasonable request.

