## Supplementary Information for "Stable isotope labeling kinetics of neurofilament light *in vitro* and *in vivo*"

**Supplementary Methods**

**Supplementary Table 1.** Cell lines used in this study

**Supplementary Table 2**. Oligonucleotides used for RT-qPCR

**Supplementary Table 3.** Antibodies used for immunocytochemistry

**Supplementary Table 4.** Monitored peptides and ion transitions for profiling and quantitation of unlabelled and labelled NfL by IP-MS/MS and yeast enolase for monitoring mass spectrometry performance

**Supplementary Table 5.** Kinetic measurements in conditioned media from iPSC-neurons

**Supplementary Figure 1.** Solubility profile of NfL in brain tissue

**Supplementary Figure 2.** Characterization of the cell lines used in this study

**Supplementary Figure 3.** Data related to NfL SILK kinetics *in vitro*

**Supplementary Figure 4**. Single peptide curves from NfL-SILK in iPSC-derived neurons from three non-degenerative donors

**Supplementary Figure 5.** Technical characterization of the NfL peptide-level IP assay

**Supplementary References**

This supplementary material has been provided by the authors to give readers additional information about their work.

### Supplementary Methods

**UCL TANGLES SILK study recruitment criteria**

Participants who fulfilled clinical criteria for corticobasal degeneration (CBD; Armstrong et al., 2013), progressive supranuclear palsy (PSP) and behavioral variant frontotemporal dementia (bvFTD; by confirmed genetic mutations in MAPT), and had given prior consent for participation in research studies were recruited from specialist clinics at University College London Hospital and through Join Dementia Research.

All donors undergoing brain donation or their next of kin gave written informed consent at registration. Queen Square Brain Bank protocols and part of the study involving donated brain tissue have been approved by the NHS Health Research Authority, Ethics Committee London-Central (REC reference 23/LO/0044).

**UCL Normal Pressure Hydrocephalus (NPH) SILK study recruitment criteria**

The UCL NPH SILK study was approved by the Bloomsbury ethics committee and all individuals provided informed written consent. Individuals with suspected idiopathic normal pressure hydrocephalus were recruited from the specialist hydrocephalus service at the National Hospital for Neurology and Neurosurgery, Queen Square.

**Differentiation of induced-pluripotent stem cells (iPSC) into iPSC-derived cortical neurons**

**Supplementary Table 1**. **Cell lines used in this study**

| Cell line | Control/mutation | Sex | Age of onset | Age at biopsy | APOE genotype |
|---|---|---|---|---|---|
| Ctrl1 (Sigma-Aldrich) | Cognitively normal SIGi1001-a-1 | F | - | 20-24 | 3/4 |
| Ctrl2 (Sigma-Aldrich) | Cognitively normal RBi001-a | M | - | 45-49 | 3/3 |
| Ctrl3 (Dr Tilo Kunath) | Healthy control line | M | - | 75-79 | 3/3 |

Briefly, iPSCs at 100% confluence were subject to neural induction using dual SMAD inhibition in N2B27 media (1 μM dorsomorphin and 10 μM SB431542, both TOCRIS). N2B27 media consists of 50% DMEM-F12, 50% Neurobasal supplemented with 0.5X N2 supplement, 0.5X B27 supplement, 0.5X L-glutamine, 0.5X non-essential amino acids, 0.5X penicillin/streptomycin, insulin (25U) and β-mercaptoethanol (1:1000). Cultures were passaged using Dispase. Progenitors underwent a final passage using Accutase at 35 DIV and neuronal maturation was performed in N2B27 media. Day 70 was taken as the starting point for *in vitro* NfL-SILK.

To monitor the successful differentiation into neurons, we isolated RNA and 4% paraformaldehyde-fixed cells on coverslips from cultures at different points during differentiation (usually at day 0 (iPSC), day 50, day 70, day 100, day >100 (between 100-120 days)).

<u>*Quantitative PCR*</u>: RNA was extracted using phenol:chloroform extraction of TriZol (Sigma) and isolated using the Monarch Nucleic Acid Purification Kit according to manufacturer's instructions. RNA concentration was quantified using a NanoDrop spectrophotometer and 1 μg of total RNA was reverse transcribed to cDNA using Superscript IV (Thermo Fisher) and random hexamers. Gene expression was quantified using ~10 ng cDNA template and Power SYBER Green PCR Master Mix (2X) (Thermo Fisher) using an MX300P real time PCR cycler (Agilent). The relative changes in mRNA expression were calculated using the comparative Ct method and were normalised to the housekeeping gene glyceraldehyde-3-phosphate (GAPDH). The list of primers used in this study can be found in **Supplementary Table 2**.

<u>*Immunocytochemistry*</u>: Coverslips with cells were fixed in 4% paraformaldehyde for 15 minutes and stored in phosphate buffered saline (PBS) until immunostaining was performed. Cells were permeabilized in 0.3% triton-X-100 in PBS for 30 minutes, then blocked in 5% bovine serum albumin (BSA) in PBS for 1 hour. Following

three washes in PBS, cells were incubated overnight in primary antibodies (see **Supplementary Table 3**). Cells were washed thrice in PBS and incubated in secondary antibodies (AlexaFluor 488, 568, 594, 647; Thermo Fisher Scientific) for 1 hour, light protected. DAPI was added as a nuclear counterstain at 0.2 ug/ml. Mounted coverslips were imaged using a Zeiss LSM microscope and Leica LAS software.

**Supplementary Table 2. Oligonucleotides used for RT-qPCR**

| Gene name | Protein encoded | Forward Sequence (5'-3') | Reverse Sequence (5'-3') |
|---|---|---|---|
| *GAPDH* | Glyceraldehyde-3-Phosphate Dehydrogenase | CGCTCTCTGCTCCTCCTGTT | CCATGGTGTCTGAGCGATGT |
| *OCT3/4* | Octamer-binding transcription factor 3/4 | TTCTGGCGCCGGTTACAGAACCA | GACAACAATGAAAATCTTCAGGAGA |
| *TBR1* | T-box brain transcription factor 1 | ATCCACAGACCCCCTCACTAG | ATCCACAGACCCCCTCACTAG |
| *TUBB3* | Tubulin beta-3 chain | CATGGACAGTGTCCGCTCAG | CAGGCAGTCGCAGTTTTCAC |
| *NEFL* | Neurofilament light chain | TGAGGAATGGTTCAAGAG | TGATCGTGTCCTGCATAG |

**Supplementary Table 3. Antibodies used for immunocytochemistry**

| Antigen | Provider | Catalog number | Dilution |
|---|---|---|---|
| NANOG | Cell Signalling Technologies | 4903 | 1:200 |
| SSEA4 | Biolegend | 330402 | 1:200 |
| NEFL | (T.400.5) Thermo Fisher | MA5-14981 | 1:200 |
| TBR1 | Abcam | ab31940 | 1:500 |
| TUBB3 (TUJ1) | Biolegend | 802001 | 1:1000 |
| Ki67 | BD Pharmingen | 550609 | 1:50 |
| MAP2 | Abcam | ab5392 | 1:500 |
| Pax6 | Biolegend | 901301 | 1:100 |

**Lactate dehydrogenase (LDH) assay**

Media from cultured cells was collected and diluted in LDH Storage Buffer (200 mM Tris-HCl pH 7.3, 10% Glycerol, 1% BSA). The dilution factor was determined by finding the linear range of the LDH positive control following titration. Samples were equilibrated at room temperature before 25 µL of each sample was added to a 96-well opaque-walled assay plate in duplicate wells. The following controls were also set up in duplicate: no-cell control (containing only culture medium) and maximum LDH release control (obtained from cells treated with 10% Triton X-100). To each well, 25 µL of LDH Detection Reagent (25 µL LDH Detection Enzyme and 0.12 µL Reductase Substrate, prepared for all samples) was added before a 1-hour incubation period. Absorbance at 450 nm was measured using a Tecan Spark 10M plate reader.

LDH release was used as a proxy of cell membrane permeability due to cytotoxicity. The results were reported as percentages relative to the positive control following the formula:

LDH release (%) = (Experimental LDH release – Medium background) /

(Experimental LDH release positive control – Medium background)

**NfL-SILK assay development and characterization**

We previously developed two targeted mass spectrometry assays for NfL; peptide-level immunoprecipitation-tandem mass spectrometry (IP-MS/MS) for targeted quantitation of a NfL Coil 2b peptide, and protein-level immunoprecipitation-mass spectrometry (IP-MS) assay for quantitation of peptides from Coils 1a, 1b, 2b and tail subdomain b of NfL[1]. Both assays were adapted for SILK by addition of monitored ion transitions for $^{13}C_6$-leucine labelled peptides (**Supplementary Table 3**).

We assessed the sensitivity of the peptide-level IP method to detect isotopically-labelled NfL using custom TLEIEACR (Coil 2b, [residues 316-323]) AQUA QuantPro peptides spiked into an artificial CSF (aCSF) matrix at different tracer ($^{13}C_6$-leucine peptide) to tracee (unlabelled peptide) ratios (TTR), with unlabelled NfL pitched at the expected average concentration in elderly healthy controls (600 pg/mL)[2]. The assay limit of detection (LOD) and limit of quantitation (LOQ) were calculated by linear regression of the calibration curve, and using the following equations:

$$LOD = (3.3 * \text{Std error of regression})/\text{slope})$$

$$LOQ = (10 * \text{Std error of regression})/\text{slope})$$

The limit of detection (LoD) and limit of quantitation (LoQ) were determined to be 0.08% TTR and 0.25% TTR respectively (**Supplementary Figure 5**).

**NfL-SILK assay *in vitro***

Sample collection

Cells were washed once with PBS and collected into 1.5 mL microfuge tubes using a cell scrapper, followed by centrifugation at 300 x g for 10 minutes at 4°C. After removing PBS, cell pellets were snap frozen on dry ice and stored at -80C until the entire experiment was completed. Conditioned media from a 12-well plate (1 mL/well) was pooled in 15 mL falcon tubes and centrifuged at 300 x g for 10 minutes at room temperature to remove cell debris. Supernatant was transferred to 15 mL falcon tubes and stored at -80C.

*Intracellular NfL-SILK*: Cells were lysed using RIPA buffer (10 mM Tris-HCl pH 7.5, 140 mM NaCl, 0.5 mM EGTA, 1 mM EDTA, 1 Triton X-100, 0.1 sodium deoxycholate, 0.1 SDS) freshly supplemented with protease inhibitor (complete Mini, EDTA-free cocktail tablets, Roche). Cell membranes were mechanically disrupted using a syringe and sonicated using a sonicator Ultrasonic Processor XL for 3 cycles of 4s at 40 amplification. Lysates were pre-cleared at 16,000 x g for 10 minutes at 4°C and the soluble fraction (supernatant) collected in a separate LoBind (Eppendorf) tube.

Protein precipitation was performed with three volumes of ice-cold acetone for 16 hours at -20°C. Precipitated protein pellets were resolubilized in 40 µL digest buffer (100 mM Tris-HCl pH 8, 6 M Urea, 2 M Thiourea, 2 ASB-14) (spiked with 1 ng of a heavy labelled [13C, 15N -Arg/Lys] recombinant NfL standard (Promise Proteomics, France) in the profiling experiments in Figure 1) and 150 ng of yeast enolase as internal control. Proteins were reduced by incubation with 1,4-dithioerythritol (90 µg/sample) for 1 hour with shaking before being alkylated by addition of iodoacetamide (216 µg/sample) and left to incubate in the dark for 1 hour. Samples were diluted with addition of ultrapure grade water (331 µL) and digested with addition of MS grade Trypsin/Lys C (Promega) to a final concentration of 2 µg/ml for 16 hours at 37°C with shaking. Trypsin was quenched by the addition of an equal volume of 0.2% formic acid (FA) in $H_2O$. Resultant tryptic peptides of NfL were isolated and cleaned up via SPE (C18 Bond Elut, Agilent Technologies). Stationary phase was wetted with 70% ACN, 0.1% FA (1 mL) and re-equilibrated with 0.1% FA (3 additions of 1 mL). NfL peptides were loaded to Toptips via centrifugation (500 rpm, 5 minutes) and washed by adding 0.1% FA (three additions of 1 mL) and centrifugation. Peptides were eluted by adding 70% ACN, 0.1% FA (two additions of 250 µL) and centrifugation. Cleaned peptide extracts were concentrated by evaporation of eluent *in vacuo* and reconstituted in 40 µL 3% acetonitrile with 0.1% FA. 10 µL were typically used for each multiple reaction monitoring (MRM) NfL assay.

*Extracellular NfL-SILK*: Sample mastermix stock containing 25X NP-40 (for a final 1X), 125 mM guanidine hydrochloride (for a final 5 mM), 25X protease inhibitor cocktail (for a final 1X) in PBS was added to conditioned media (10 mL) and incubated under rotation at 4°C for 30 minutes. Samples were concentrated using 15 mL Amicon filters with a 10kDa cut-off at 4500 rpm for 30 minutes at room temperature. This step concentrated the media down to ca 300-500 µL (approx. 20X). *Immunoprecipitation:* Concentrated media was incubated with mouse IgG2a-coupled M-270 Epoxy Dynabeads™ for 1 hour at 4°C to exclude unspecific bindings to the beads. In parallel, M-270 Epoxy Dynabeads™ coupled to anti-NfL HJ30.13, HJ30.4 and HJ30.11 antibodies, or mouse IgG as negative control, (approx. 9 µg of antibodies/sample) were blocked using horse myoglobulin in PBS (1 mg/ml) for 1 hour at 4°C. Concentrated, pre-cleared media was incubated with anti-NfL/anti-IgG coupled beads overnight at 4°C. Beads were then washed three times with 1 mL of 25 mM triethylammonium bicarbonate (TEABC). NfL-coupled beads were suspended in 40 µL digest buffer (containing recombinant heavy labelled NfL in the profiling experiments in Figure 1) and 75 ng yeast enolase for on-bead reduction, alkylation, digestion, SPE clean up and resuspension prior to UPLC-MS/MS analysis as described above.

**NfL SILK assay *in vivo***

*UCL TANGLES SILK study:* Briefly, individuals received a $^{13}C_6$-leucine infusion intravenously at 4 mg/patient kg/hr for 16 hours. When infusion stopped, CSF was collected by lumbar puncture (LP) across 5 visits, which fell on the following number of days post-infusion: day 4 (LP1), day 8 (LP2), between days 13-15 (LP3), between days 53-67 (LP4) and between days 112-162 (LP5). CSF was centrifuged at 1000 xg or 10 minutes at 4°C, aliquoted into protein low protein bind tubes (MAXYmum Recovery, Axygen) and stored at -80°C within 1 hour of collection.

*NPH SILK study:* Briefly, individuals received a 13C6-leucine infusion intravenously at 2 mg/patient kg/hr for between 3 and 16 hours prior to either diagnostic lumbar drain insertion or ventriculoperitoneal shunt insertion. A full thickness cortical biopsy was collected during VP shunt insertion as previously described[3].

**Soluble and insoluble fractions of *post-mortem* brain tissue**

Briefly, 0.27 – 0.57 gram of grey matter from frontal cortex tissue was homogenized in 10 volumes (v/w) of lysis buffer (10 mM Tris–HCl, pH 7.4, 800 mM NaCl, 1 mM EDTA, 2 mM DTT, 10% sucrose) using a probe sonicator (TissueRuptor, QIAGEN) on ice until completely homogenized. The starting brain homogenate was centrifuged at 16000 x g for 20 minutes at 4°C. The crude supernatant was treated with N-lauroylsarcosinate ("sarkosyl", 1% [w/v] final concentration) and shaken at room temperature for 1 hour. The supernatant was then centrifuged at 100000 x g for 1 hour at 4°C. The sarkosyl-insoluble pellet was resuspended in 300 µL washing buffer (10 mM Tris–HCl, pH 7.4, 800 mM NaCl, 5 mM EDTA, 2 mM DTT, 10% sucrose) and centrifuged at 16000 x g for 30 minutes at 4°C. After centrifugation, the supernatant was further centrifuged at 100000 x g (Optima MAX, Beckman Coulter) for 1 hour at 4°C. The supernatant was kept in protein LoBind tubes (Eppendorf) as the sarkosyl-soluble fraction for immunoprecipitation of NfL. The pellet was resolubilized in 70% FA using a probe sonicator and dried down by evaporation of eluent *in vacuo.*

**NfL-SILK quantitation**

Acquired data was imported into Skyline software (MacCoss Lab, University of Washington) for peak picking and integration, and QC checks including retention time stability, quantifier/qualifier ion ratios and signal/noise measures. To calculate NfL tracer-to-tracee ratio (TTR), labeled peak areas were divided by unlabeled peak areas and represented as percentages. NfL kinetic curves were plotted in GraphPad Prism and RStudio. The half-life of NfL was calculated by fitting each peptide-level data into a one-phase decay model. For calculating the average half-life of NfL, the estimated half-lives for single peptides were used. Estimates where a double-sided confidence interval could not be reported were considered unreliable and excluded. Peptides containing two leucine resides in their sequence were normalized following the formula:

$$TTR\_corrected\_for\_2\_leu\_double\_label = TTR/(TTR + 0.5)$$

The fractional synthesis rate (FSR) was calculated using the standard formula:

$$FSR = (Et2 - Et1)NfL/(t2 - t1) / \text{Precursor } E$$

where $(Et2 - Et1)NfL/(t2 - t1)$ was defined as the slope of the linear regression from 3 to 24 days of labeling divided by the leucine enrichment in media (at 50 mol, this equals 1). The fractional clearance rate (FCR) was calculated by fitting the slope of the natural logarithm of the "clearance" portion of the labeled NfL curve (i.e, the chase) according to the formula:

$$FCR = \ln(\text{labeled NfL}/\text{unlabeled NfL}) / (t2 - t1)$$

**Leucine enrichment in plasma**

To quantitate labelled leucine enrichment in plasma, a hydrophilic interaction liquid chromatography – tandem mass spectrometry (HILIC-MS/MS) assay was adapted from Prinsen et al[4] to measure $^{13}C_6$-/$^{12}C_6$-leucine ratios, with the following optimisations and adaptations:

Analysis was performed using an Acquity H-Class Ultra Performance Liquid Chromatography (UPLC) system, fitted with an ACQUITY UPLC BEH Amide column (100A; 1.7µm, 2.1 x 50mm) attached to a VanGuard UPLC BEH Amide precolumn (2.1 x 5 mm), which was coupled to a Xevo TQ-S triple quadrupole mass spectrometer operated in positive electrospray ionisation (ESI+) mode (Waters, UK). Chromatographic separation was performed over a 10-minute HILIC gradient using mobile phases A (10 mM ammonium formate in 85% ACN 0.15% FA) and B (10mM ammonium formate in ultrapure Milli-Q water 0.15% FA). The column was primed and equilibrated for 45 minutes in initial conditions (100% A at 0.4 mL/min) and kept at 35˚C. At injection (1 µL/sample), the column was kept in initial conditions for 3 minutes until a linear gradient of increasing %B began. From 3 – 3.1 minutes B was increased to 5.9%, and from 3.1 – 5 minutes was increased to 17.6%, followed by a final increase to 29.4% from 5 – 6 minutes. The flow rate was then increased to 0.6 mL/min and the column re-equilibrated in 100% A for 4 minutes. Mass spectrometer parameters were set as follows: source temperature (150˚C), capillary voltage (1.00 kV), desolvation temperature (550˚C), desolvation gas flow (1000 L/hr) and cone gas flow (150 L/hr).

Ion transitions for $^{12}C_6$-leucine (precursor: 132.102 m/z, product: 86.100 m/z), 13C6-leucine (precursor: 138.122 m/z, product: 91.139 m/z) and 13C6-,15N2-lysine internal standard (precursor: 155.127 m/z, product: 90.100 m/z) were analysed by multiple reaction monitoring (MRM) and acquired data imported into Skyline (MacCoss Lab, Seattle, USA) for processing and peak integration. Peak areas were exported into Microsoft Excel and leucine TTR calculated as molar $^{13}C_6$-leucine/$^{12}C_6$-leucine peak area ratios.

For TANGLES, the leucine enrichment in plasma was calculated as the average of leucine enrichment at plateau points (between 6-16 hours of labelling) and reported as percentage (**Table 1**). Because labelling in the NPH SILK cohort was variable (dependent on surgery timings), not all subjects showed a plateau in the plasma leucine enrichment. Leucine enrichment data from the two NPH subjects with *ex-vivo* brain biopsies is taken from one subject in the NPH SILK cohort that showed plateau and reported as percentage (**Table 1**).

**Graphics**
All schematics were produced using BioRender (www.biorender.com) under an active licensed agreement.

**Supplementary Table 4. Monitored peptides and ion transitions for profiling and quantitation of unlabelled and labelled NfL by IP-MS/MS and yeast enolase for monitoring mass spectrometry performance.**

| Structural domain | Amino acids | Peptide sequence | Precursor ion (m/z) | Precursor charge (z) | Product ion (type) | Product ion (m/z) |
|---|---|---|---|---|---|---|
| Head | 31-37 | SGYSTAR | 371.1799 | 2 | y4+ | 434.2358 |
| | | | | | y5+ | 597.2991 |
| | | SGYSTAR [ISTD] | 376.1841 | 2 | y4+ | 444.2440 |
| | | | | | y5+ | 607.3074 |
| Coil 1a | 92-100 | AQLQDLNDR | 536.7727 | 2 | y7+ | 873.4425 |
| | | | | | y5+ | 632.2998 |
| | | AQLQDLNDR [SILK] | 539.7828 | 2 | y7+ | 879.4626 |
| | | | | | y5+ | 638.32 |
| | | AQLQDLNDR [SILK] | | | y7+ | 879.4626 |
| | | | | | y5+ | 632.2998 |
| | | AQLQDLNDR [SILK] | 542.7928 | 2 | y7+ | 885.4827 |
| | | | | | y5+ | 638.32 |
| | | AQLQDLNDR [ISTD] | 541.7769 | 2 | y7+ | 883.4507 |
| | | | | | y5+ | 642.3081 |
| | 101-107 | FASFIER | 435.2294 | 2 | y5+ | 651.3461 |
| | | | | | y6+ | 722.3832 |
| | | FASFIER [ISTD] | 440.2336 | 2 | y5+ | 661.3543 |
| | | | | | y6+ | 732.3914 |
| Coil 1b | 117-126 | VLEAELLVLR | 577.8606 | 2 | y8+ | 942.5619 |
| | | | | | y7+ | 813.5193 |
| | | VLEAELLVLR [SILK] | 583.8809 | 2 | y8+ | 948.582 |
| | | | | | y7+ | 819.5394 |
| | | VLEAELLVLR [ISTD] | 582.8649 | 2 | y8+ | 952.5701 |
| | | | | | y7+ | 823.5275 |
| | 137-144 | ALYEQEIR | 511.2693 | 2 | y6+ | 837.4101 |
| | | | | | y4+ | 545.3042 |
| | | ALYEQEIR [SILK] | 514.2793 | 2 | y6+ | 837.4101 |
| | | | | | y4+ | 545.3042 |
| | | ALYEQEIR [ISTD] | 516.2734 | 2 | y6+ | 847.4184 |
| | | | | | y4+ | 555.3125 |
| | 148-157 | LAAEDATNEK | 531.2591 | 2 | y8+ | 877.3898 |
| | | | | | y6+ | 677.3101 |
| | | LAAEDATNEK [SILK] | 534.2692 | 2 | y8+ | 877.3898 |
| | | | | | y6+ | 677.3101 |
| | | LAAEDATNEK [ISTD] | 535.2662 | 2 | y8+ | 535.2662 |
| | | | | | y6+ | 685.3243 |
| | 158-164 | QALQGER | 401.2143 | 2 | y3+ | 361.183 |
| | | | | | y4+ | 489.2416 |

| **Structural domain** (continued) | **Amino acids** (continued) | **Peptide sequence** (continued) | **Precursor ion (m/z)** (continued) | **Precursor charge (z)** (continued) | **Product ion (type)** (continued) | **Product ion (m/z)** (continued) |
|---|---|---|---|---|---|---|
| | | QALQGER [SILK] | 404.2244 | 2 | y3+ | 361.183 |
| | | | | | y4+ | 489.2416 |
| | | QALQGER [ISTD] | 406.2184 | 2 | y3+ | 371.1913 |
| | | | | | y4+ | 499.2499 |
| | 165-172 | EGLEETLR | 473.7456 | 2 | y4+ | 518.2933 |
| | | | | | y3+ | 389.2507 |
| | | EGLEETLR [SILK] | 476.7557 | 2 | y4+ | 518.2933 |
| | | | | | y3+ | 395.2708 |
| | | EGLEETLR [SILK] | 479.7658 | 2 | y4+ | 524.3134 |
| | | | | | y3+ | 395.2708 |
| | | EGLEETLR [ISTD] | 478.7498 | 2 | y4+ | 528.3016 |
| | | | | | y3+ | 399.2590 |
| | 178-185 | YEEEVLSR | 512.7509 | 2 | y6+ | 732.3886 |
| | | | | | y5+ | 603.3461 |
| | | YEEEVLSR [SILK] | 515.761 | 2 | y6+ | 738.4088 |
| | | | | | y5+ | 609.3662 |
| | | YEEEVLSR [ISTD] | 517.7551 | 2 | y6+ | 742.3969 |
| | | | | | y5+ | 613.3543 |
| | 198-206 | GADEAALAR | 437.2249 | 2 | y5+ | 501.3144 |
| | | | | | y4+ | 430.2772 |
| | | GADEAALAR [SILK] | 440.2349 | 2 | y5+ | 507.3345 |
| | | | | | y4+ | 436.2974 |
| | | GADEAALAR [ISTD] | 442.2290 | 2 | y5+ | 511.3226 |
| | | | | | y4+ | 440.2855 |
| Coil 2b | 284-293 | FTVLTESAAK | 533.7926 | 2 | y8+ | 818.4618 |
| | | | | | y7+ | 719.3934 |
| | | FTVLTESAAK [SILK] | 536.8027 | 2 | y8+ | 824.4819 |
| | | | | | y7+ | 725.4135 |
| | | FTVLTESAAK [ISTD] | 537.7997 | 2 | y8+ | 826.4760 |
| | | | | | y7+ | 727.4076 |
| | 316-323 | TLEIEACR | 496.2475 | 2 | y6+ | 777.356 |
| | | | | | y5+ | 648.3134 |
| | | TLEIEACR [SILK] | 499.2576 | 2 | y6+ | 777.356 |
| | | | | | y5+ | 648.3134 |
| | | TLEIEACR [ISTD] | 501.2516 | 2 | y6+ | 787.3642 |
| | | | | | y5+ | 658.3216 |
| | 324-331 | GMNEALEK | 446.2157 | 2 | y6+ | 703.3621 |
| | | | | | y4+ | 460.2766 |
| | | GMNEALEK [SILK] | 449.2257 | 2 | y6+ | 709.3822 |
| | | | | | y4+ | 466.2967 |
| | | GMNEALEK [ISTD] | 450.2228 | 2 | y6+ | 711.3763 |
| | | | | | y4+ | 468.2908 |

| **Structural domain** (continued) | **Amino acids** (continued) | **Peptide sequence** (continued) | **Precursor ion (m/z)** (continued) | **Precursor charge (z)** (continued) | **Product ion (type)** (continued) | **Product ion (m/z)** (continued) |
|---|---|---|---|---|---|---|
| Tail Subdomain A | 422-437 | SAYGGLQTSSYLMSTR | 861.4118 | 2 | y9+ | 1045.498 |
| | | | | | y8+ | 944.4506 |
| | | SAYGGLQTSSYLMSTR [SILK] | 864.4219 | 2 | y9+ | 1045.4983 |
| | | | | | y8+ | 944.4506 |
| | | SAYGGLQTSSYLMSTR [ISTD] | 866.4159 | 2 | y9+ | 1055.5065 |
| | | | | | y8+ | 954.4589 |
| Tail Subdomain B | 530-540 | VEGAGEEQAAK | 544.7646 | 2 | y9+ | 860.4108 |
| | | | | | y7+ | 732.3523 |
| | | VEGAGEEQAAK [ISTD] | 548.7717 | 2 | y9+ | 868.4250 |
| | | | | | y7+ | 740.3665 |
| | 16-28 | GNPTVEVELTTEK | 708.8645 | 2 | y11++ | 623.3323 |
| | | | | | y8+ | 948.4884 |

Abbreviations: ISTD, internal standard

For each NfL peptide, ion transitions are provided for endogenous NfL (unlabelled), $^{13}C_6$-leucine labelled NfL [SILK] and the $^{13}C_6$, $^{15}N$-labelled Arg/Lys NfL internal standard [ISTD].

Quantifier and qualifier product ions are listed for each peptide, with the quantifier listed first.

For stable isotope labelled peptides ([SILK] or [ISTD]), the labelled amino acid is in bold and underlined (ie. L for $^{13}C_6$-Leucine or K/R for $^{13}C_6$-, $^{15}N$-labelled Lysine/Arginine respectively).

**Supplementary Table 5. Kinetic measurements in conditioned media from iPSC-neurons**

| FRACTIONAL SYNTHESIS RATE (Extracellular) | | | | | | | | | | | | |
|---|---|---|---|---|---|---|---|---|---|---|---|---|
| | Ctrl1 | | | | | | Ctrl2 | | | Ctrl3 | | |
| **Peptide** | **Amino acids** | **n1** | **n2** | **n3** | **n4** | **Mean FSR (%/d) ± SD** | **n1** | **n2** | **Mean FSR (%/d) ± SD** | **n1** | **n2** | **Mean FSR (%/d) ± SD** |
| AQLQDLNDR | 92-100 | 2.4057 | 2.5862 | NA | 1.6374 | 2.209767 | 1.2277 | 0.992 | 1.10985 | 2.20 | 1.64 | 1.92 ± 0.39 |
| ALYEQEIR | 137-144 | 2.698 | 2.8696 | NA | 1.8588 | 2.475467 | 1.3648 | 0.5864 | 0.9756 | 2.71 | 2.14 | 2.42 ± 0.40 |
| LAAEDATNEK | 148-157 | 2.885 | 2.7799 | NA | 2.0544 | 2.5731 | 1.3677 | 1.0414 | 1.20455 | 2.44 | 2.22 | 2.33 ± 0.15 |
| FTVLTESAAK | 284-293 | 3.1673 | 3.4784 | NA | 1.9353 | 2.860333 | 1.2051 | 1.1979 | 1.2015 | 3.63 | 1.24 | 2.43 ± 1.69 |
| TLEIEACR | 316-323 | 2.8992 | 3.1666 | NA | 2.5434 | 2.869733 | 1.579 | 1.2254 | 1.4022 | 3.16 | 2.19 | 2.67 ± 0.68 |
| GMNEALEK | 324-331 | 2.9341 | 3.538 | NA | 2.5314 | 3.001167 | 1.5316 | 1.2575 | 1.39455 | 3.17 | 2.34 | 2.75 ± 0.59 |
| | Mean all | | | | | **2.75** ± 0.30 | | | **1.21** ± 0.16 | | | **2.42** ± 0.29 |
| **FRACTIONAL CLEARANCE RATE (Extracellular)** | | | | | | | | | | | | |
| **Peptide** | **Amino acids** | **n1** | **n2** | **n3** | **n4** | **Mean FCR (%/d) ± SD** | **n1** | **n2** | **Mean FCR (%/d) ± SD** | **n1** | **n2** | **Mean FCR (%/d) ± SD** |
| AQLQDLNDR | 92-100 | 4.74 | 6.01 | NA | 4.03 | 4.93 | 2.36 | 0.15 | 1.2550 | 4.32 | 1.79 | 3.06 ± 1.79 |
| ALYEQEIR | 137-144 | 4.80 | 3.95 | NA | 9.95 | 6.23 | 2.92 | 0.99 | 1.9550 | 8.56 | 0.34 | 4.45 ± 5.81 |
| LAAEDATNEK | 148-157 | 4.08 | 4.22 | NA | 5.38 | 4.56 | 3.49 | 1.10 | 2.2950 | 5.25 | 1.07 | 3.16 ± 2.96 |
| FTVLTESAAK | 284-293 | 2.19 | 3.29 | NA | 4.80 | 3.43 | 1.76 | 1.07 | 1.4150 | 5.58 | 2.92 | 4.25 ± 1.88 |
| TLEIEACR | 316-323 | 4.05 | 5.22 | NA | 6.00 | 5.09 | 3.04 | 1.79 | 2.4150 | 6.92 | 3.12 | 5.02 ± 2.69 |
| GMNEALEK | 324-331 | 4.52 | 5.20 | NA | 5.93 | 5.22 | 3.13 | 1.31 | 2.2200 | 7.61 | 5.54 | 6.58 ± 1.46 |
| | Mean all | | | | | **4.90** ± 0.92 | | | **1.92** ± 0.48 | | | **4.42** ± 1.30 |

SD: standard deviation; FSR: fractional synthesis rate (represented as percentage per day ± SD); FCR: fractional clearance rate (represented as percentage per day ± SD); NA: not available; n: induction number; amino acids in the consensus sequence of NfL as per UniProt entry P07196.

**Supplementary Figure 1. Solubility profile of NfL in brain tissue**

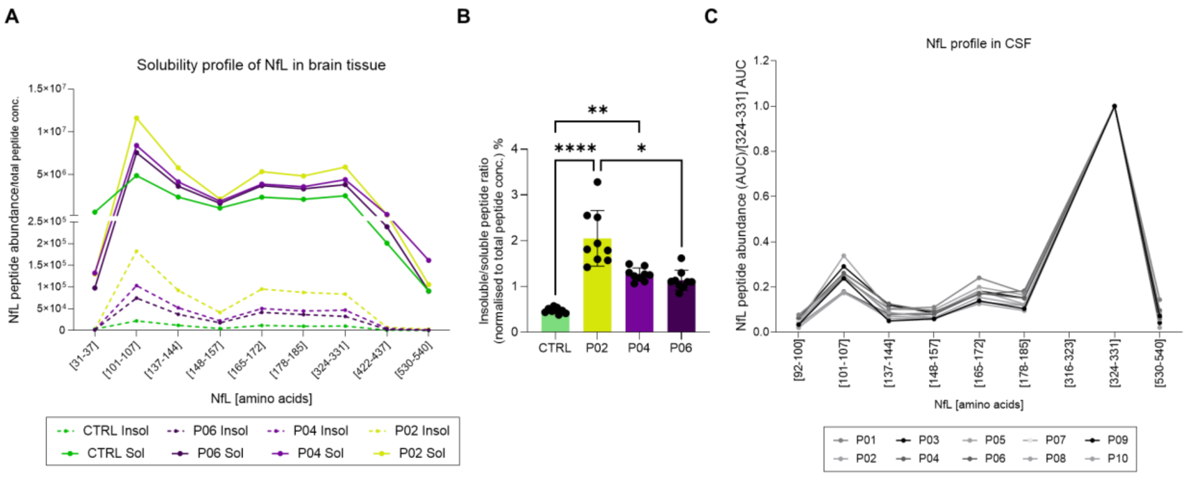


**A)** Peptide recovery across the NfL sequence after immunoprecipitation normalised to total peptide concentration in the samples. Recovery of NfL was lower in insoluble compared to soluble fractions. Peptide [31-37] abundance was higher in CTRL compared to P02, P04 and P06. **B)** The peptide abundances normalised to total peptide content in the sarkosyl-insoluble fraction ("insoluble") were divided by their abundance in the sarkosyl-soluble ("soluble") fractions to calculate the ratio of insoluble NfL peptides across cases, shown as a percentage (%). Each dot represents a peptide (including peptides within residues [31-37], [101-107], [137-144], [148-157], [165-172], [178-185], [324-331], [422-437], [530-540] of the NfL amino acid sequence). Results are displayed as mean ± SD. Kruskal-Wallis test with Dunn's multiple comparison's test. Comparisons were made between all groups; whenever not shown, the testing showed no significant differences. **C)** CSF recovery of NfL proteotypic peptides in individual participants from the TANGLES study. **$p \leq 0.01$; ****$p \leq 0.0001$. CTRL = healthy control. Insol = Insoluble. Sol = Soluble.

**Supplementary Figure 2. Characterization of the cell lines used in this study**

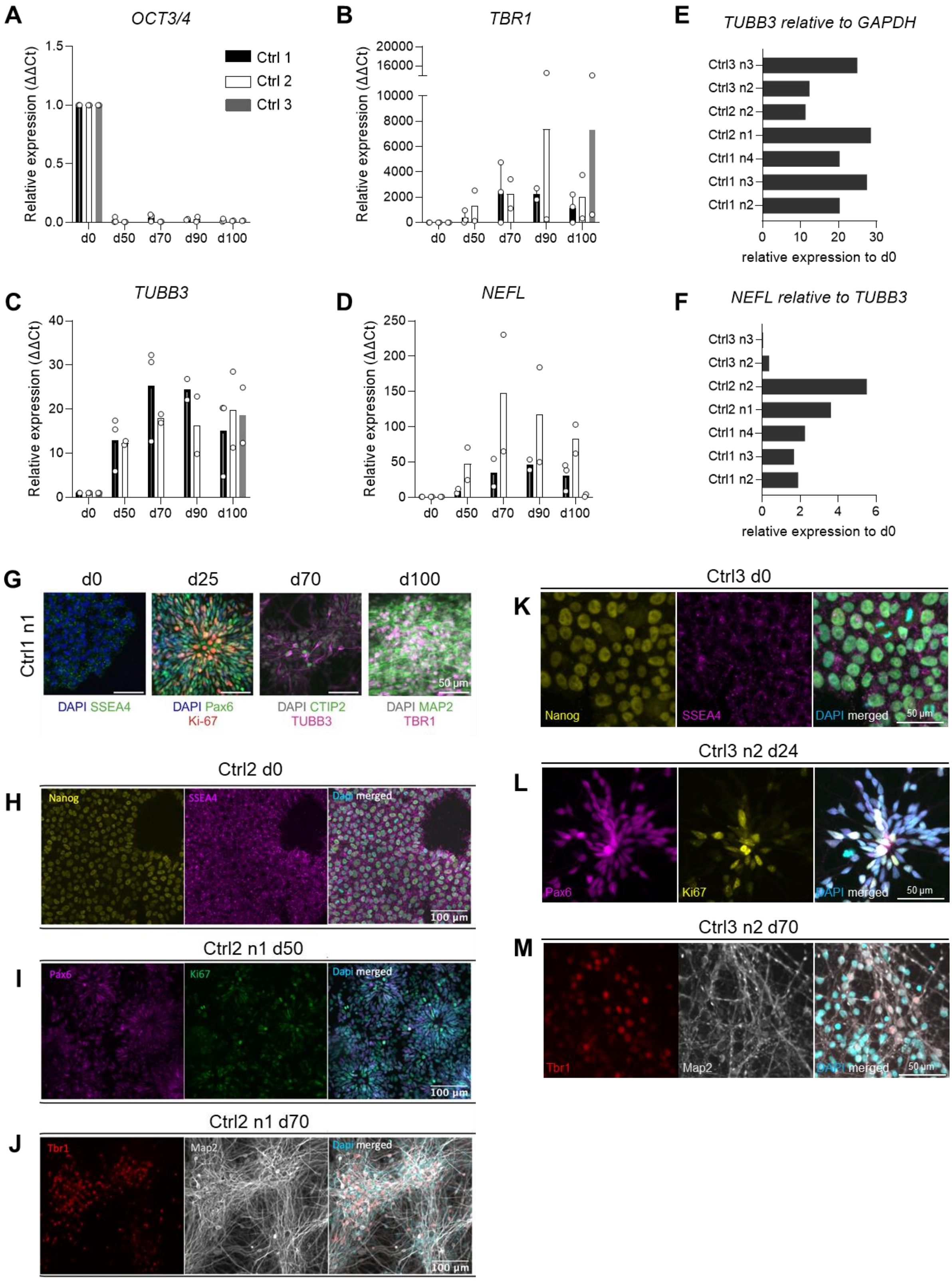


**A)** Relative levels of the pluripotency gene Octamer-binding transcription factor 3/4 (*OCT3/4*), **B)** T-box brain transcription factor 1 (*TBR1*), **C)** Beta III tubulin (*TUBB3*), **D)** Neurofilament light (*NEFL)* in the different iPSC lines used. Ctrl1 and Ctrl2 include data at d0, d50, d70, d90, d100. Ctrl3 include data at d0 and d100 only (due to a lower yield of neurons, only 100 DIV was prioritized for this analysis). **E)** *TUBB3* expression relative to the housekeeping gene *GAPDH* in the independent inductions used in the study. **F)** *NEFL* expression relative to

*TUBB3* in the independent inductions used in the study. **G)** Widefield fluorescence microscopy images of Ctrl1 iPSC stained for the pluripotency markers Nanog and Stage-specific embryonic-antigen 4 (SSEA4) (d0); paired box 6 (Pax6) and Ki67 demonstrating conversion into actively dividing Neural Precursor Cells (NPCs) at 25 DIV (d25); the deep-layer neuronal marker COUP-TF-interacting protein 2 (CTIP2) and the pan neuronal microtubule-associated protein 2 (MAP2) at 70 DIV (d70); the upper-layer neuronal marker T-box brain transcription factor 1 (TBR1) and MAP2 at 100 DIV (d100) Scale bar = 50 µm. Ctrl2 n1 cells stained for **H)** Nanog and SSEA4 in iPSCs (d0); **I)** Pax6 and Ki67 at 50 DIV (d50); **J)** TBR1 and MAP2 at 70 DIV (d70). Scale bar = 100 µm. Ctrl3 cells n3 stained for **K)** Nanog and SSEA4 in iPSCs (d0); **L)** Pax6 and Ki67 at 24 DIV (d24); **M)** TBR1 and MAP2 at 70 DIV (d70). Scale bar = 50 µm. In all immunostainings, DAPI was used as a nuclear counterstain and is only shown in the merged images.

**Supplementary Figure 3. Data related to NfL-SILK kinetics *in vitro*.**

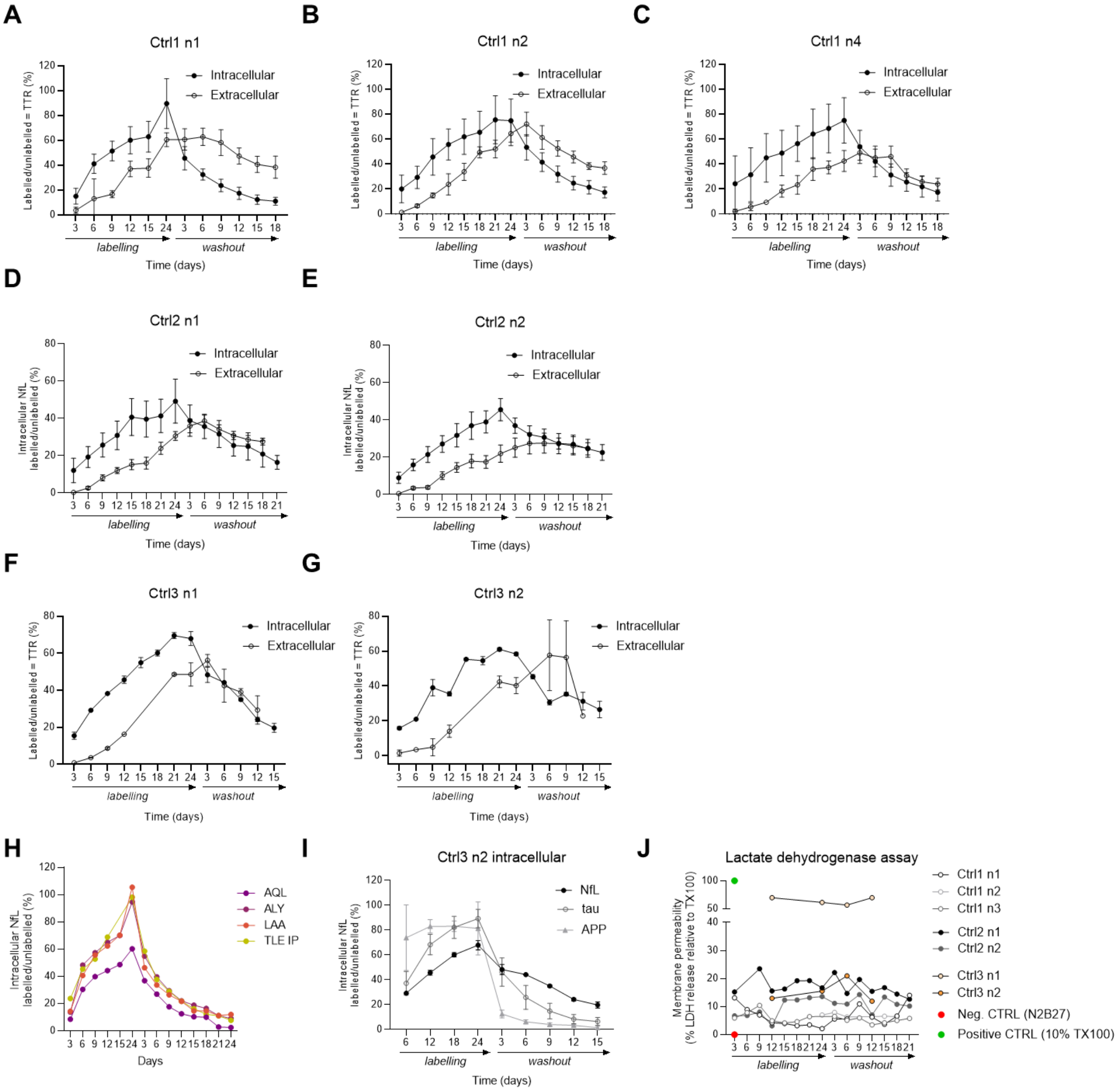


Kinetics in cell lysate ("intracellular") and cell media ("extracellular") in three independent inductions of Ctrl1 **(A, B, C)**, two inductions of Ctrl2 **(D, E)** and two inductions of Ctrl3 **(F, G)** cells. **H)** Kinetic curves of proteotypic peptides in NfL (depicted by the first three amino acids of the peptide in 1-letter code) compared to the kinetic profile obtained when performing peptide-level IP with anti-TLEIEACR antibodies ("TLE IP") in Ctrl1 cells (induction n1). **I)** Kinetic profiles of NfL, tau and amyloid precursor protein (APP) obtained in Ctrl3 cells (induction n2). **J)** Lactate dehydrogenase release in cell media was measured to assess the levels of cell cytotoxicity in the cultures over the course of the labelling and washout periods of the experiments. N2B27 only was used as negative control. Cells treated with 10% Triton X-100 were used as positive control. Data are shown as measured LDH release divided by maximum LDH release (elicited by treatment of the positive control sample with 10% Triton X-100 [TX100]).

**Supplementary Figure 4. Single peptide curves from NfL-SILK in iPSC-derived neurons from three non-degenerative donors**.

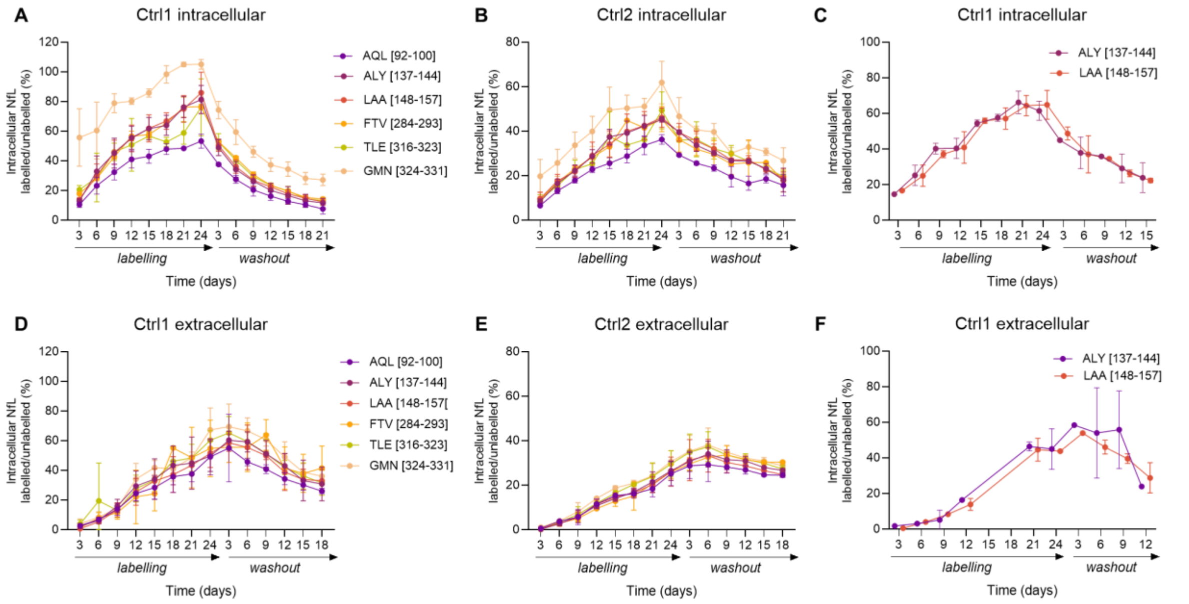


**A)** Intracellular Ctrl1 results (n=3 independent inductions). **B)** Intracellular Ctrl2 results (n=2 independent inductions). **C)** Intracellular Ctrl3 intracellular results (n= 2 independent inductions). **D)** Extracellular Ctrl1 results (n=3 independent inductions). **E)** Extracellular Ctrl2 results (n=2 independent inductions). **F)** Extracellular Ctrl3 Extracellular results (n= 2 independent inductions). Datapoints represent the mean tracer-to-tracee ratio (TTR) of the peptides at any given timepoint from all inductions ± SD. Peptides are depicted by the first three amino acids of the peptide in 1-letter code, followed by the peptides' first and last residues within the NfL amino acid sequence in brackets.

**Supplementary Figure 5. Technical characterization of the NfL peptide-level IP assay.**

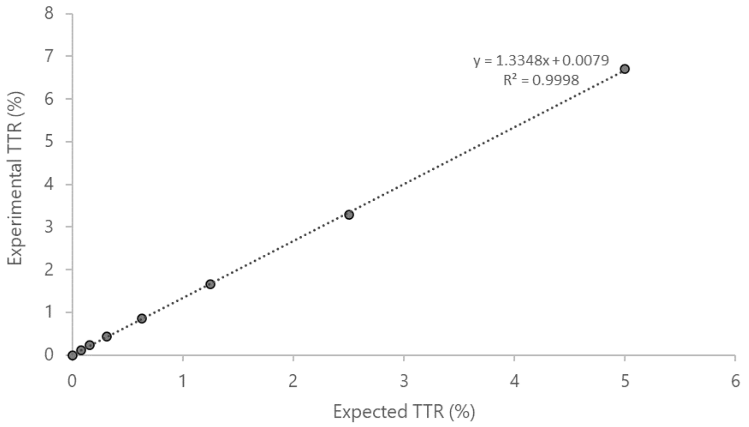


Custom AQUA QuantPro peptides for the Coil 2b peptide (TLEIEACR) were spiked in artificial CSF (aCSF) to assess the sensitivity of the assay as TTR %, where unlabeled NfL was pitched at 600 pg/mL. From characterization, limit of detection (LoD) and limit of quantitation (LoQ) for SILK TTR% in aCSF was determined to be 0.08% and 0.25% respectively.
